# Pilot program for test-based SARS-CoV-2 screening and environmental monitoring in an urban public school district

**DOI:** 10.1101/2021.04.14.21255036

**Authors:** John Crowe, Andy T. Schnaubelt, Scott Schmidt-Bonne, Kathleen Angell, Julia Bai, Teresa Eske, Molly Nicklin, Catherine Pratt, Bailey White, Brodie Crotts-Hannibal, Nicholas Staffend, Vicki Herrera, Jeramie Cobb, Jennifer Conner, Julie Carstens, Jonell Tempero, Lori Bouda, Matthew Ray, James V. Lawler, W. Scott Campbell, John-Martin Lowe, Joshua Santarpia, Shannon Bartelt-Hunt, Michael Wiley, David Brett-Major, Cheryl Logan, M. Jana Broadhurst

## Abstract

**Importance:** Scalable programs for school-based SARS-CoV-2 testing and surveillance are needed to guide in-person learning practices and inform risk assessments in K-12 settings.

**Objectives:** To characterize SARS-CoV-2 infections in staff and students in an urban public school setting and evaluate test-based strategies to support ongoing risk assessment and mitigation for K-12 in-person learning.

**Design, Setting, and Participants:** The pilot program engaged three schools for weekly saliva PCR testing of staff and students participating in in-person learning over a 5-week period. Wastewater, air, and surface samples were collected weekly and tested for SARS-CoV-2 RNA to determine surrogacy for case detection and interrogate transmission risk of in-building activities.

**Main Outcomes and Measures:** SARS-CoV-2 detection in saliva and environmental samples and risk factors for SARS-CoV-2 infection.

**Results:** 2,885 supervised self-collected saliva samples were tested from 773 asymptomatic staff and students during November and December, 2020. 46 cases (22 students, 24 staff) were detected, representing a 5.8- and 2.5-fold increase in case detection rates among students and staff, respectively, compared to conventional reporting mechanisms. SARS-CoV-2 RNA was detected in wastewater samples from all pilot schools, as well as in air samples collected from two choir rooms. Sequencing of 21 viral genomes in saliva specimens demonstrated minimal clustering associated with one school. Geographic analysis of SARS-CoV-2 cases reported district-wide demonstrated higher community risk in zip codes proximal to the pilot schools.

**Conclusions and Relevance:** Weekly screening of asymptomatic staff and students by saliva PCR testing dramatically increased SARS-CoV-2 case detection in an urban public-school setting, exceeding infection rates reported at the county level. Experiences differed among schools, and virus sequencing and geographic analyses suggest a dynamic interplay of school-based and community-derived transmission risk. Environmental testing for SARS-CoV-2 RNA in air and surface samples enabled real-time risk assessment of in-school activities and allowed for interventions in choir classes. Wastewater testing demonstrated the utility of school building-level SARS-CoV-2 surveillance. Collectively, these findings provide insight into the performance and community value of test-based SARS-CoV-2 screening and surveillance strategies in the K-12 educational setting.

**KEY POINTS:** *Question:* Do test-based programs reduce SARS-CoV-2 risk in K-12 schools?

*Findings:* Weekly school-based saliva PCR testing at three urban public schools doubled case detection among staff and students over symptom-based strategies, exceeding county-level case rates. SARS-CoV-2 was detected in school wastewater samples each week, as well as air and surface samples related to choir classrooms.

*Meaning:* Routine SARS-CoV-2 testing removes infected staff and students from school who are not identified through conventional case detection. With rigorous infection control and environmental monitoring, this helps mitigate risk during school operations. Furthermore, screening in K-12 schools may provide insight into disease burdens of undertested communities.

## Introduction

During the fall and winter of 2020 most Northern Hemisphere countries experienced their most severe COVID-19 epidemic waves to date. While many factors may have contributed to the acceleration of SARS-CoV-2 transmission in the fall, the return of children and young adults to congregate school settings was temporally associated with the initiation of increased community transmission. Overall, direct data demonstrating transmission of SARS-CoV-2 among school-aged children within communities remain limited.^1^ Many experts and public health authorities have urged communities to resume full-time in-person learning, citing the absence of school-associated case clusters and the lower rate of laboratory-confirmed SARS-CoV-2 diagnosis among school-age children versus adults.^2,3,4^ Yet, despite lower rates of confirmed infections in children and adolescents, serological surveys suggest current symptom-based testing and tracing likely miss a large proportion of disease, especially in children and adolescents.^5^ Data through November 2020 from the U.S. Centers for Disease Control and Prevention (CDC) COVID-19 serological survey revealed that persons aged 0-17 had the highest proportion of anti-SARS-CoV-2 antibodies of any age group in 23 of 28 (82%) states providing data.^6^ Outbreaks of COVID-19 in schools have been well-documented,^7^ including a large outbreak in a school in Israel that resulted in student and staff attack rates of 13% and 17% respectively.^8^

To promote safe school-environments, the World Health Organization (WHO), U.S. CDC and other health policy centers have recommended combining interventions such as physical distancing, the wearing of face masks, enhanced environmental cleaning, and use of physical barriers to reduce the risk of SARS-CoV-2 transmission.^9,10^ Many U.S. school systems have chosen to use virtual or hybrid education models, rather than full-density in-person schooling. In developing risk mitigation plans, public health professionals and school administrators have been forced to weigh incomplete data characterizing COVD-19 school risks with harms of suspending in-person educational services. To make better decisions, school officials require an improved understanding of COVID-19 burden in the households relevant to schools and greater characterization of in-school transmission risks. Additionally, regular testing to rapidly identify students or staff who are a risk for transmitting COVID-19 may significantly reduce the risk of outbreaks centered in schools, enable more effective learning environments, and ultimately slow community transmission. As SARS-CoV-2 variants expand in the U.S. and the length of protective immunity remains uncertain, these needs persist into the 2021-2 school year as well as future respiratory disease emergencies.

The University of Nebraska Medical Center (UNMC) and Omaha Public Schools (OPS) partnered to launch the **PRO**active **TE**sting for **C**ommunity **T**ransmission of **S**ARS-COV-2 (OPS PROTECTS) pilot project in November 2020 to demonstrate the feasibility of a school-based COVID-19 testing program. The OPS PROTECTS pilot investigated the integration of individual case detection through saliva testing with school-level wastewater monitoring and in-building air and surface sampling for SARS-CoV-2 RNA – approaches that have not yet been described in the K-12 educational setting. The pilot sought to comprehensively assess SARS-CoV-2 infection and transmission risk in three pilot schools to demonstrate the feasibility of expanding to additional schools during the COVID-19 pandemic.

## Methods

### Pilot program setting

The Omaha Public School district is comprised of 82 primary and secondary schools, roughly 20 programs, and more than 53,000 students and 9,200 school-based staff. Two middle schools and one high school in a medically underserved sector of the Omaha metropolitan area were selected to participate in the pilot program with the goal of maximizing the benefit of early program implementation. During the Fall 2020 semester, all schools in the district were operating under a hybrid instructional model with alternating cohorts for remote and optional in-person learning with 50-60% of students opting for in-person learning. District-wide policies to mitigate SARS-CoV-2 transmission risk include the utilization of in-building face coverings, six-foot social distancing, and heightened handwashing and cleaning protocols. The five-week pilot program took place from November 9-December 11, 2020.

### Saliva SARS-CoV-2 PCR testing for asymptomatic case detection

Electronic registration and consent for saliva testing was performed with the Nebraska University Laboratory Information Reporting Tool (NULirt) software platform. Staff participation was mandated by the school district while student participation was optional. Supervised self-collections of saliva were performed at mobile collection stations in each school twice weekly to accommodate alternating cohorts of students present for in-person learning at either the beginning of the week (i.e., students whose last names began with A through K) or at the end of the week (i.e., students whose last name began with L though Z). Trained volunteers supervised collections according to current CDC guidance.^11^ Staff testing began during the first week of the pilot and student testing began during the second week. Saliva was collected using half-length plastic straws (S.P. Richards Co, Atlanta, GA) and 1.5mL collection tubes (Eppendorf Co, Hamburg, Germany).

Saliva SARS-CoV-2 testing was performed by the CLIA-certified Emerging Pathogens Laboratory at UNMC using a multiplex, qualitative, real-time, reverse transcription (RT)-PCR assay adapted from the SalivaDirect™ protocol.^12,13^ Via NUlirt, test results were securely linked to the electronic medical record, public health databases, and individual results were securely emailed to participants. Individuals with a positive test result were isolated from in-person activities for ten days and contacted by a healthcare provider to offer access to counseling and clinical services.

### School facility wastewater testing for SARS-CoV-2 RNA

Wastewater grab samples were collected twice weekly from near-source outflow locations that conveyed wastewater from each of the three pilot schools. Grabs samples were collected between (11:00-13:00) from manholes adjacent to the buildings on school grounds. At each sampling event, 250 mL of raw wastewater was collected in a sterile polypropylene container for SARS-CoV-2 molecular analysis and 250 mL of raw wastewater was collected in a non-sterile polypropylene container for conventional wastewater analyses. All samples were held at 4°C after collection and were transported to UNMC or UNL laboratories within 2 hours of collection, where they were held at −20°C until RT-PCR analysis for SARS-CoV-2 using the IDT 2019-nCoV RUO kit (IDT, 10006713).

### School building air and surface testing for SARS-CoV-2 RNA

Weekly air and surface sampling was performed in each pilot school at five sites per school that were assessed to be at higher risk for virus aerosol exposure, including band and choir rooms, cafeterias, language teaching classrooms, high-traffic hallways, restrooms, and school entry areas. Air samples were collected using AirAnswers (Inspirotec1, Chicago, Il., USA) air samplers^14^ which collect particles by electrostatic precipitation. This sampler does not use a pump or blower to pull air into the sampler, minimizing noise disruption in the school environment. Electrostatic precipitation has been used in the collection of a variety of airborne particulate^15,16^ including viruses.^17^ Surface samples were collected on doors leading into each of the air sampling spaces. Extracted samples were analyzed by RT-PCR for the SARS-CoV-2 E gene as previously described.^18^

### Demographic and statistical analyses

Metadata associated with SARS-CoV-2 test results included collection location and date, school, status as student or staff, zip code of residence, grade in school for students, main teaching activity or occupation role for staff, and participation in band and choir. Non-pilot program cases among students were identified through school-level designated absentee reports. Staff non-pilot program cases were identified through self-report to the school district. Background community COVID-19 rates were obtained via the Douglas County Health Department, Nebraska on-line dashboard.^21^

Additional information on methods is available in the supplement.

## Results

### Saliva testing program metrics and participant demographics

Registration and consent numbers for staff and students over the five-week program period are shown in eFigure 1. 2,885 saliva samples were tested from 773 participants [2,163 tests from 455 (96%) school-based staff and 722 tests from 315 (12%) students enrolled for in-person learning]. Both students and staff were ethnically diverse, and >89% of student households were eligible for financial assistance (Table 1). 340 volunteer shifts were filled over the course of the program to supervise saliva collection (approximately 60 person-hours per school each week). Almost all (99.4%) saliva samples met laboratory acceptability criteria and yielded valid test results with an average turnaround time of less than 5 hours from receipt in the laboratory to reporting results.

**Table 1:** Student and staff demographics in pilot schools.

|  |  | School A<br>(Grades 9-12) |  | School B<br>(Grades 6-8) |  | School C<br>(Grades 5-8) |  | All Program<br>Schools |  |
| --- | --- | --- | --- | --- | --- | --- | --- | --- | --- |
|  |  | No. | % | No. | % | No. | % | No. | % |
| **Students eligible for free or reduced-price lunch** |  |  |  |  |  |  |  |  |  |
|  | Participants | 98 | 70.5 | 61 | 85.9 | 85 | 81.0 | 244 | 77.5 |
|  | Total | 1546 | 86.3 | 1081 | 91.3 | 1075 | 91.3 | 3702 | 89.1 |
| **Race/Ethnicity, Students** |  |  |  |  |  |  |  |  |  |
| American Indian<br>or Alaska Native | Participants | 1 | 0.7 | 1 | 1.4 | 0 | 0 | 2 | 0.6 |
|  | Total | 16 | 0.9 | 9 | 0.8 | 6 | 0.5 | 31 | 0.7 |
| Asian | Participants | 0 | 0 | 0 | 0 | 0 | 0 | 0 | 0 |
|  | Total | 28 | 1.6 | 5 | 0.4 | 1 | 0.1 | 34 | 0.8 |
| Black or African<br>American | Participants | 10 | 7.2 | 5 | 7.0 | 5 | 4.8 | 20 | 6.4 |
|  | Total | 225 | 12.6 | 87 | 7.3 | 31 | 2.6 | 343 | 8.3 |
| Hispanic | Participants | 77 | 55.4 | 43 | 60.6 | 81 | 77.1 | 201 | 63.8 |
|  | Total | 1220 | 68.1 | 898 | 75.8 | 2036 | 88.0 | 3154 | 76.0 |
| Native Hawaiian<br>or Pac. Islander | Participants | 0 | 0 | 0 | 0 | 0 | 0 | 0 | 0 |
|  | Total | 2 | 0.1 | 0 | 0 | 1 | 0.1 | 3 | 0.1 |
| White | Participants | 42 | 30.2 | 16 | 22.5 | 17 | 16.2 | 75 | 23.8 |
|  | Total | 242 | 13.5 | 158 | 13.3 | 87 | 7.4 | 487 | 11.7 |
| Two or more<br>races | Participants | 9 | 6.5 | 6 | 8.5 | 2 | 1.9 | 17 | 5.4 |
|  | Total | 59 | 3.3 | 27 | 2.3 | 15 | 1.3 | 101 | 2.4 |
| **Race/Ethnicity, Staff<sup>a</sup>** |  |  |  |  |  |  |  |  |  |
| American Indian<br>or Alaska Native | Participants | 2 | 1.1 | 0 | 0 | 0 | 0 | 2 | 0.4 |
| Asian | Participants | 7 | 3.9 | 1 | 0.8 | 3 | 2.1 | 11 | 2.4 |
| Black or African<br>American | Participants | 12 | 6.7 | 9 | 6.8 | 4 | 2.8 | 25 | 5.5 |
| Hispanic | Participants | 31 | 17.4 | 16 | 12.1 | 36 | 24.8 | 83 | 18.2 |
| Native Hawaiian<br>or Pac. Islander | Participants | 1 | 0.6 | 0 | 0 | 0 | 0 | 1 | 0.2 |
| White | Participants | 118 | 66.3 | 101 | 76.5 | 93 | 64.1 | 312 | 68.6 |
| Other race | Participants | 1 | 0.6 | 1 | 0.8 | 5 | 3.5 | 7 | 1.5 |
| Not provided | Participants | 6 | 3.4 | 4 | 3.0 | 4 | 2.8 | 14 | 3.1 |

<sup>a</sup>Due to mandatory staff participation in the testing program, participants represent >90% of total staff across the pilot schools

### SARS-CoV-2 asymptomatic case detection by weekly saliva testing

Forty-six total cases (24 staff, 22 students) were detected through weekly saliva PCR testing of asymptomatic individuals. The pilot program more than doubled cases identified among staff, and nearly doubled cases identified among students (eFigure 2). Cumulative case rates detected by saliva testing among pilot program participants substantially exceeded case rates detected by conventional reporting mechanisms among students and staff registered for in-person school activities over the same time period at the pilot schools (students: 70/1000 vs 12/1000, staff: 53/1000 vs 21/1000; Figure 1). Weekly case detection rates by saliva PCR across all pilot schools ranged from 10 to 52 cases per 1000 (1 to 5.2%) students and 8 to 17 cases per 1000 (0.8 to 1.7%) staff (Figure 2). The majority of staff cases were detected within a week interval from their last negative test, while half of student cases were detected at two to three weeks from their last negative test indicating less consistent student participation (eFigure 3). Fifteen of seventeen (88%) PCR positive individuals who participated in re-testing tested negative on their first test following a 10-day isolation period.

**Figure 1:**
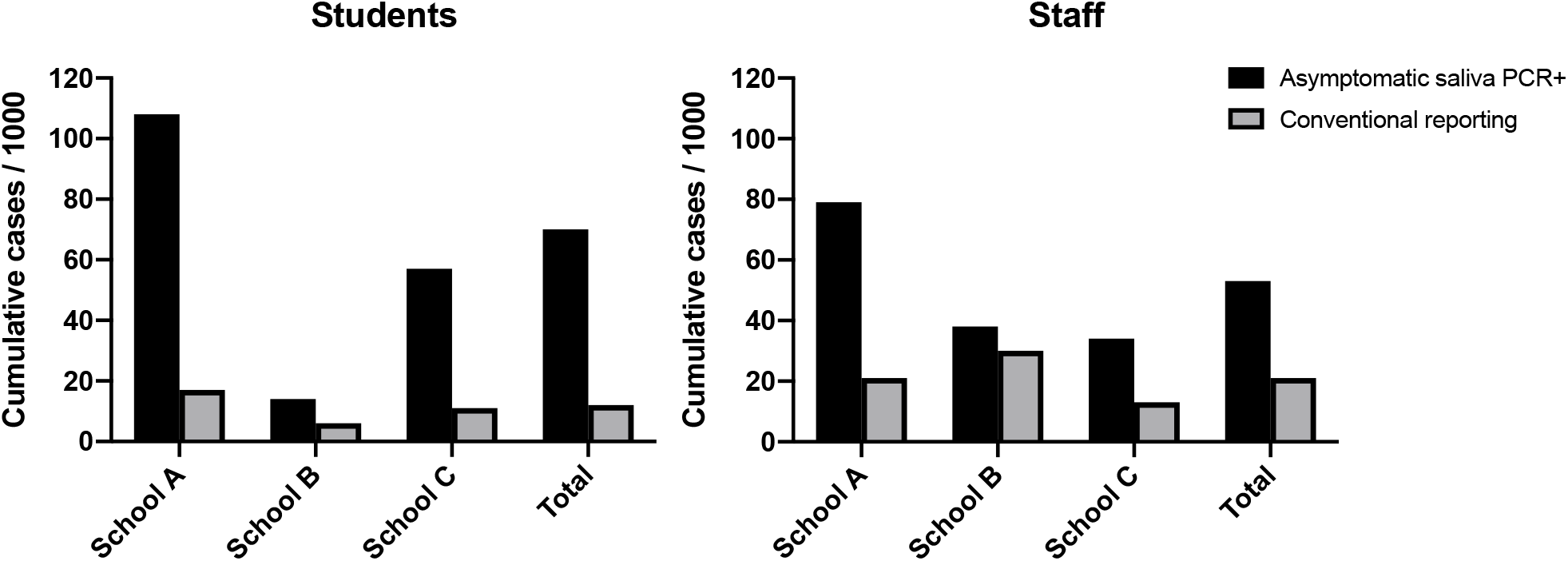
Cumulative SARS-CoV-2 case rates detected by weekly saliva PCR compared to conventional reporting during the pilot program period. For asymptomatic saliva PCR, rates are among pilot program participants; for conventional reporting, rates are among individuals participating in in-person school activities.

**Figure 2:**
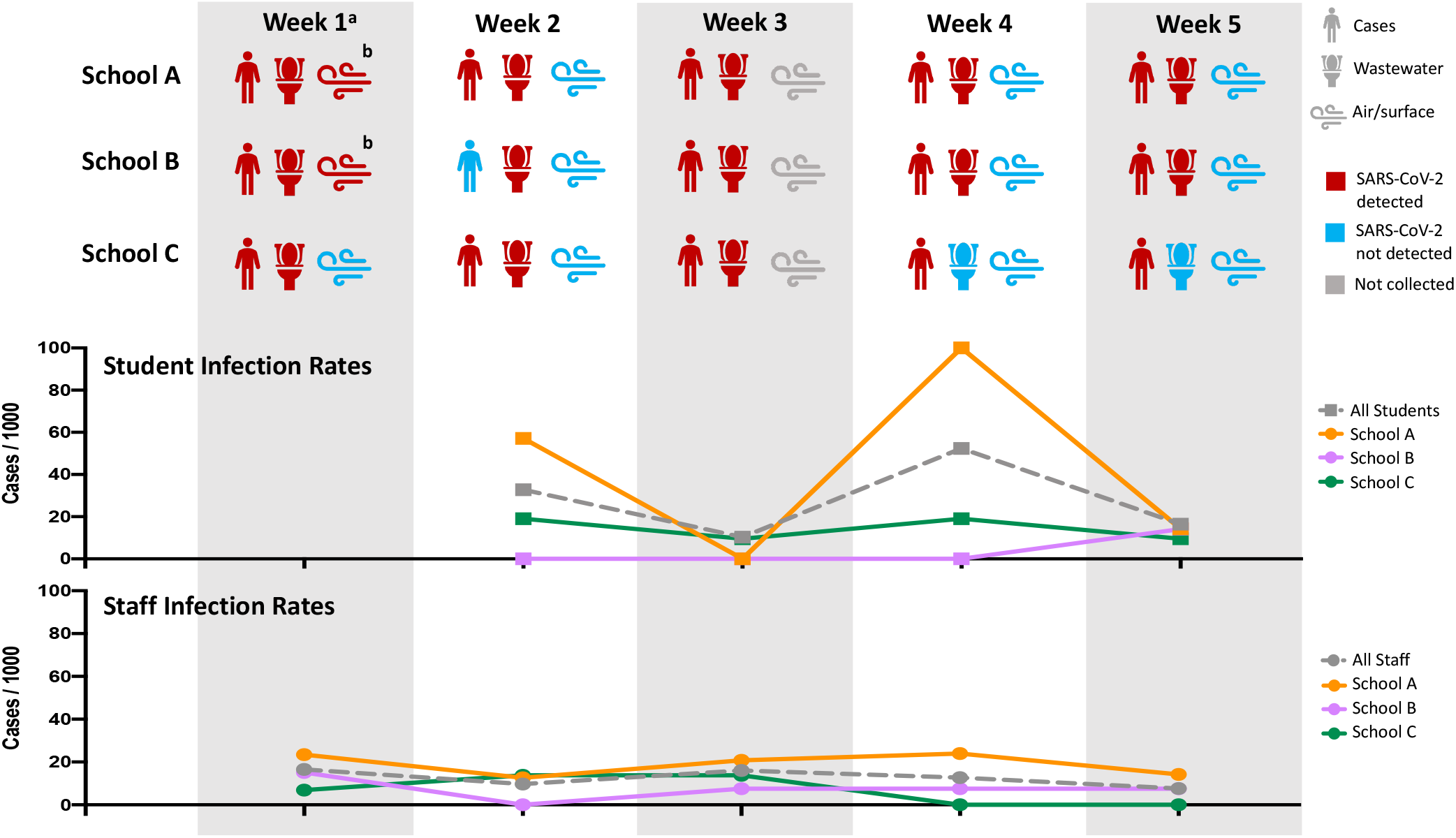
SARS-CoV-2 detected by saliva testing and environmental surveillance methods. Cases represent those detected by weekly saliva PCR testing. ^a^ Staff saliva testing began during Week 1; student saliva testing began during Week 2 ^b^ Air and surface samples positive for SARS-CoV-2 RNA collected in choir rooms

### SARS-CoV-2 RNA detection in school facility wastewater and in-building environmental samples

SARS-CoV-2 RNA was detected by RT-PCR in wastewater grab samples from School A and School B facilities during all five weeks of the pilot, and at School C for the first three weeks of the pilot (Figure 2). Weekly wastewater testing detected SARS-CoV-2 in 12/14 (86%) individual school-weeks for which SARS-CoV-2 was detected in students or staff by saliva testing.

In-building air and surface samples were collected from five sampling sites in each pilot school during four weeks of the pilot program (60 total paired samples). A subset of air samples (2/60; 3.3%) and surface samples (1/60; 1.7%) tested positive for SARS-CoV-2 RNA by RT-PCR. The positive air samples were collected from School A and School B choir rooms, and the positive surface swab was collected in the School B choir room (Figure 2; eTable 1).

### Virus whole-genome sequencing of SARS-CoV-2-positive saliva samples

SARS-CoV-2 genomes from positive saliva samples were sequenced to investigate transmission chains within cases identified by our testing program. We obtained high-coverage viral genomes from 21 of 46 cases and generated phylogenetic trees using the available Nebraska SARS-CoV-2 genomes from GISAID (177genomes). Most (17/ 21) SARS-CoV-2 genomes did not cluster together but were widely spread across the tree, indicating that these cases were the result of separate transmission chains (Figure 3). We observed two clustering events with samples from School A (highlighted in Figure 3), between two staff (CSSCH003 and CSSCH016, identical genomes) and two students (CSSCH033 and CSSCH042, one nucleotide difference). Clear epidemiologic links between the clustered genomes beyond school attendance was not identified; however, the timing of case identification was compatible with linked transmission events.

**Figure 3:**
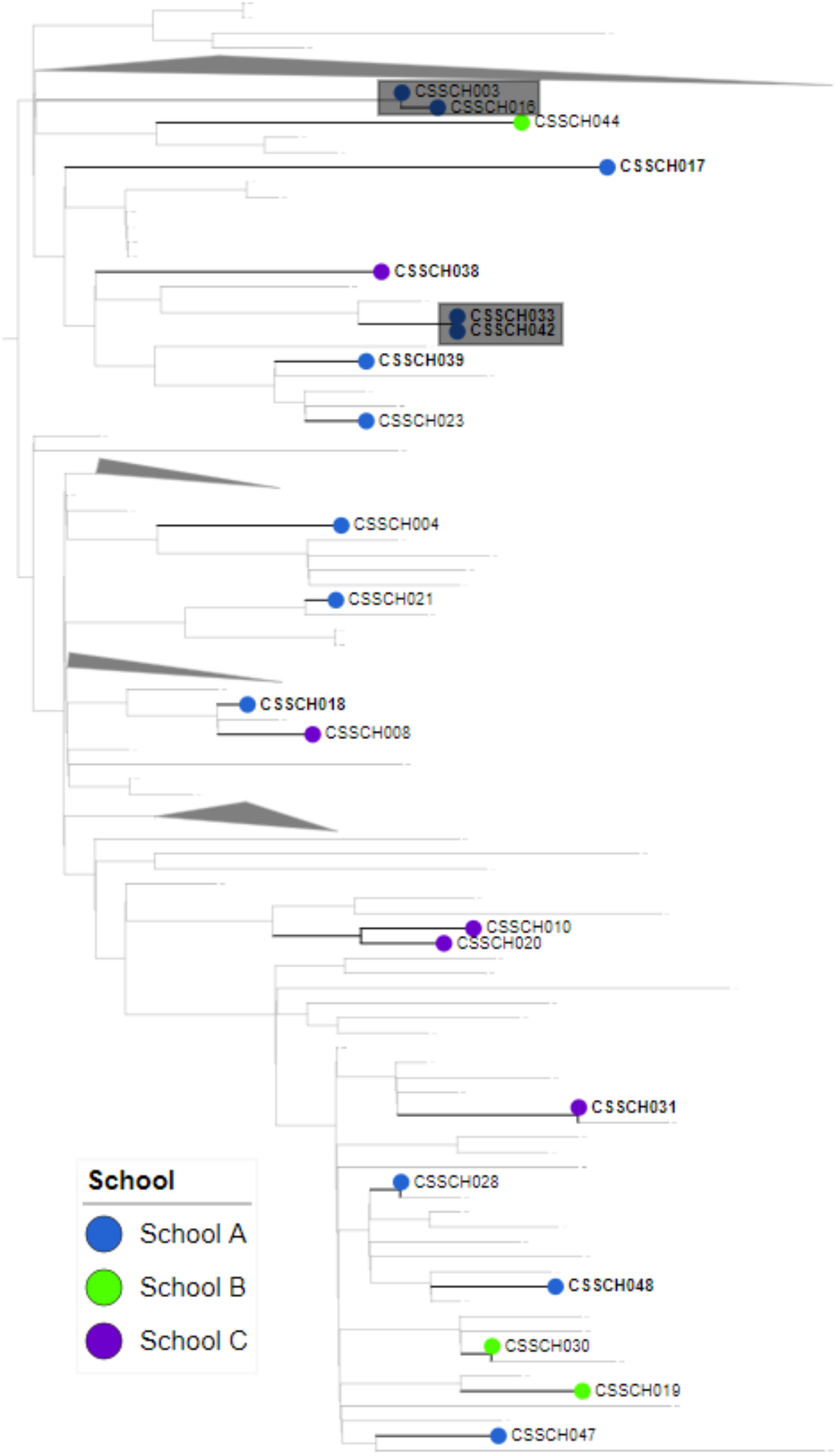
Phylogenetic analysis of SARS-CoV-2 sequences from saliva specimens collected at pilot schools. Clustered samples are shaded in grey.

### School-based risk factors for SARS-CoV-2 infection

Participation in school activities (choir and band), school attended, school grade, and staff position were evaluated as risk factors for SARS-CoV-2 infection as detected by saliva PCR testing during the pilot program (eTable 1). School attended was a significant determinant for positive test results in logistic regression models (p = 0.0063). School A (high school) students and staff were more likely than School B and C (middle schools) to test positive [3.3 (p=0.009) and 2.2 (p=0.03), respectively). Schools B and C case rates were similar.

Students in choir were 2.8 times as likely to test positive for SARS-CoV-2 than other students when adjusting for school attended, though with borderline statistical significance (p=0.052); unadjusted significance was less (p=0.19). Serial case counts among students participating in choir were assessed following the positive air samples in choir rooms during week one of the pilot (eFigure 4). Amplification of case counts in serial weeks was not observed.

Broadly, the category of staff position was not significantly associated with SARS-CoV-2 testing results. However, business teachers in the pilot (n=7) were 28.5 times as likely to test positive for SARS-CoV-2 than other staff (p<0.0001).

### Geographic distribution of district-wide COVID-19 cases reported among staff and students

Weekly case rates detected by saliva testing in the pilot schools exceeded county-level case rates identified through conventional testing means over the same time period by a log. We sought to better understand the role of localized community case burden in these findings. In preliminary bivariate analyses, zip code of residence did not achieve statistical significance as a factor associated with saliva SARS-CoV-2 test result for either students or staff at the pilot schools. To contextualize the pilot schools within the district, Figure 4 demonstrates the distribution of cases by zip code of residence in students and staff across the entire Omaha Public School district, either through self-report or district-managed occupational screening separate from the pilot program. Student case counts were highest in a South Omaha zip code with 66 cases during the pilot period, demonstrating higher case burden in the community sector where the pilot schools are located.

**Figure 4:**
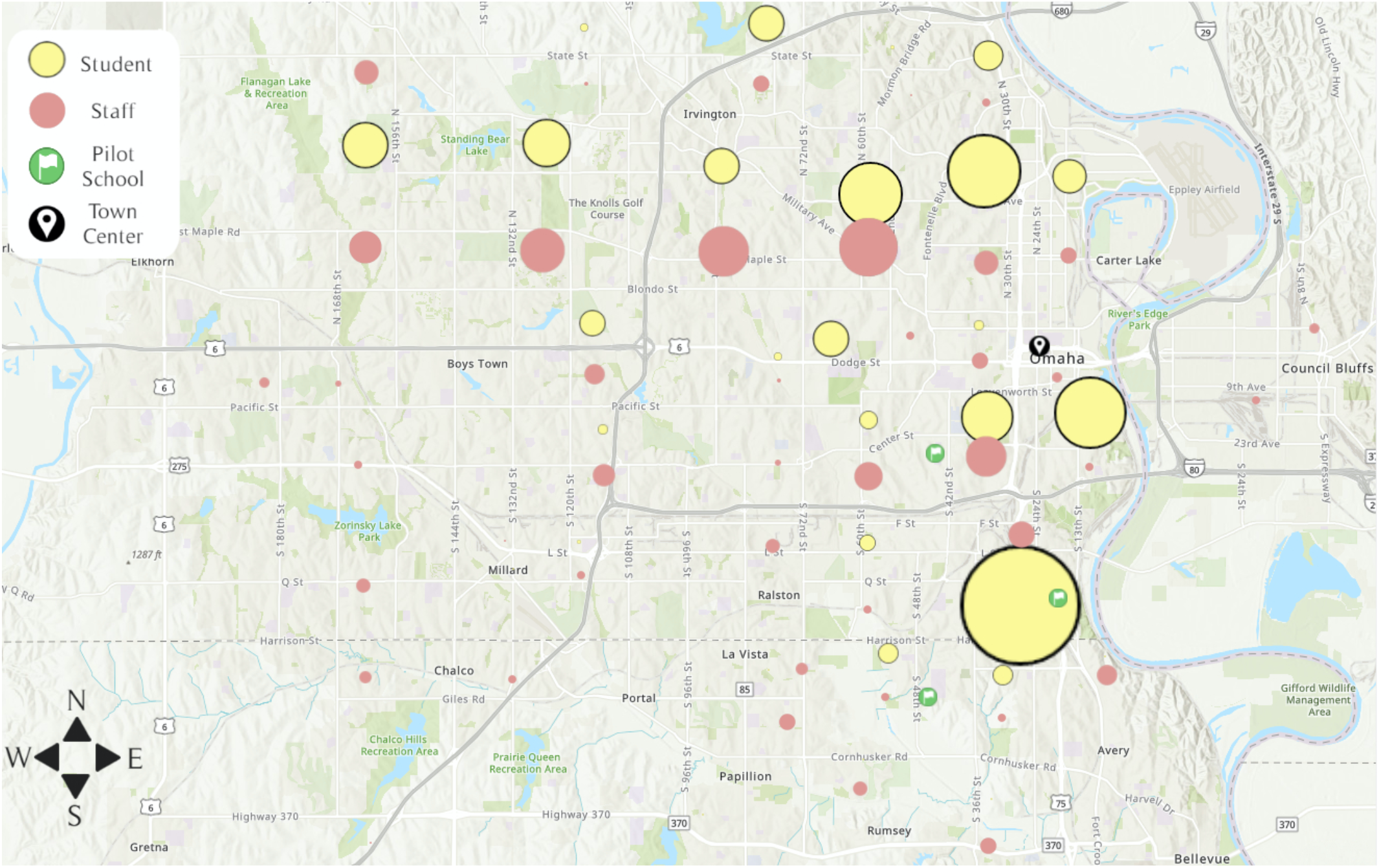
Geographic distribution of home residences linked to COVID-19 cases reported for staff and students across the school district. Circle size denotes relative number of cases reported by zip code.

## Discussion

OPS PROTECTS represents a feasible, scalable, and novel approach to integrated, test-based SARS-CoV-2 screening and environmental monitoring in a K-12 educational setting. Our pilot schools are located in a traditionally underserved area of Omaha; students at these schools predominantly identify within racial and ethnic minority groups and qualify for financial assistance. Students and staff living in zip codes near these schools experienced a heavy burden of COVID-19 cases as reported in school district-wide case tracking. Weekly saliva PCR markedly increased case finding and the removal of potential opportunities for viral transmission. Additionally, when compared to conventional reporting in our setting (passive case finding), our results suggest that as many as 9 in 10 student COVID-19 cases and 7 in 10 staff COVID-19 cases may be missed by conventional reporting mechanisms. Together with striking differences between pilot program and observed community case rates, this kind of programming can be expected to assist in mitigating school-based transmission risk through informing case isolation, contact tracing, and management of school activities and serve as a trigger to escalate community-based surveillance in underserved communities, particularly in school-aged children.

The pilot program demonstrated successful implementation of school-based SARS-CoV-2 testing through supervised self-collection of saliva. While trained volunteers from local health professions colleges supervised saliva collections during the pilot program, we found that responsibility for collection processes could be successfully transferred to school staff allowing for more sustainable and integrated operations during program expansion. Self-collected saliva samples from students and staff yielded a high rate of valid test results, indicating reliable specimen quality despite not restricting intake of food or drink prior to collection. The use of non-specialized collection materials (e.g. common plastic straws rather than saliva collection aids) and a high-throughput adaptation of the SalivaDirect™ extraction-free RT-PCR method minimized cost and supply chain challenges while also facilitating same-day reporting.

To the best of our knowledge, this study represents the first description of building-level environmental testing for SARS-CoV-2 in K-12 schools. Wastewater monitoring for SARS-CoV-2 RNA by twice-weekly grab sampling was generally consistent with the detection of SARS-CoV-2 infections by saliva testing, indicating the utility of this approach for school building-level monitoring of virus shedding. However, two wastewater collections (School C, weeks 4 and 5) yielded negative results while the schools still had persons with positive saliva tests. This highlights the need to continue to pursue optimal wastewater collection practices for population monitoring. Time-distributed composite samples collected with autosampler installations may increase sensitivity but add cost and infrastructure requirements. Wastewater monitoring by grab sampling and automated composite sampling will be further explored as distributable school surveillance tools for identification of SARS-CoV-2 transmission hotspots. Ultimately, wastewater and other environmental monitoring may allow for more resource-intensive individual screening to be prioritized, a strategy that has been successfully pursued in university housing^22,23^ and nursing home^24^ settings.

Air and surface sampling within school buildings for SARS-CoV-2 RNA provides insight into virus dispersion associated with school activities. In our pilot program, positive air and surface samples were detected in the choir rooms of two schools during the first week of the program. These findings suggest that measures to reduce virus dispersion in the school environment— including mask wearing, hand hygiene, and cleaning protocols—did not fully mitigate risk associated with choir. Choir-associated COVID-19 outbreaks have been described^25,26^ and we observed a trend toward increased risk of SARS-CoV-2 infection among students participating in choir in our pilot program. The transmission risk of other specialized school activities warrants further evaluation. For example, our initial findings suggest a higher risk for infection in teachers associated with computer labs, and we will pursue focused environmental testing in computer labs in the next phase of the program.

Although our findings confirm that incidence of SARS-CoV-2 infection in schools greatly exceeded what was observed through conventional case finding, our data do not permit firm conclusions about comparative incidence or transmission events within schools. Genomic sequencing identified potential transmission links among students and staff in two clusters at School A, but virus from saliva samples in our pilot mostly demonstrated a mix of multiple disparate transmission chains compatible with broader community transmission. Our highest weekly incidence for students (Week 4) was over seven times the reported weekly community incidence for Douglas County (week of Nov 30^th^), and the incidence in School A was 14-fold higher. However, our schools are located in communities that experienced higher incidence and where limiting testing access may accentuate under-ascertainment of cases. It is also the case that our schools were operating at one-quarter normal classroom densities so our results may underestimate the risk of in-school transmission for schools operating at more normal density. Regardless, our observations emphasize the variable risk landscape between schools and the importance of school-based testing strategies to identify school settings and activities with higher transmission risk.

Higher student participation would allow a better understanding of case rates within schools, demographic risk groups among students and staff, and correlation with wastewater and in-building environmental testing strategies. Time intervals for test conversion indicate that promoting consistent student participation, in addition to raising student consent rates, will improve timely detection and isolation of new cases. The PROTECTS pilot schools have a relatively restricted geographic distribution of residence among both students and staff. This may have attenuated the ability of the pilot to discern residence as an independent risk factor.

## Data Availability

All viral genome sequences referenced in this study have been deposited in the GISAID database.

https://www.gisaid.org

## ARTICLE INFORMATION

### Author contributions

Dr. Broadhurst had full access to all of the data in the study and takes responsibility for the integrity of the data and the accuracy of the data analysis.

#### Concept and design

John Crowe, Andy T. Schnaubelt, Scott Schmidt-Bonne, James V. Lawler, W. Scott Campbell, John-Martin Lowe, Joshua Santarpia, Michael Wiley, Shannon Bartelt-Hunt, David Brett-Major, Cheryl Logan, M. Jana Broadhurst

#### Acquisition, analysis, or interpretation of data

John Crowe, Andy T. Schnaubelt, Scott Schmidt-Bonne, Kathleen Angell, Julia Bai, Teresa Eske, Molly Nicklin, Matthew Ray, Catherine Pratt, Bailey White, Brodie Crotts-Hannibal, Nick Staffend, Vicki Herrera, Jennifer Conner, Julie Carstens, Lori Bouda, James V. Lawler, John-Martin Lowe, Joshua Santarpia, Michael Wiley, Shannon Bartelt-Hunt, David Brett-Major, Cheryl Logan, M. Jana Broadhurst

#### Drafting of the manuscript

John Crowe, Andy T. Schnaubelt, Catherine Pratt, Joshua Santarpia, Michael Wiley, Shannon Bartelt-Hunt, David Brett-Major, M. Jana Broadhurst

#### Critical revision of the manuscript for important intellectual content

James V. Lawler, John-Martin Lowe, Matthew Ray, Cheryl Logan

#### Statistical analysis

Scott Schmidt-Bonne, Teresa Eske, Molly Nicklin, Kathleen Angell, Julia Bai, David Brett-Major

#### Obtained funding

John Crowe, Cheryl Logan, M. Jana Broadhurst

#### Administrative, technical, or material support

Nick Staffend, W. Scott Campbell, Teresa Eske, Molly Nicklin

#### Supervision

John Crowe, W. Scott Campbell, John-Martin Lowe, Joshua Santarpia, Michael Wiley, Shannon Bartelt-Hunt, David Brett-Major, Cheryl Logan, M. Jana Broadhurst

### Additional Contributions

The authors would like to thank the clinical partners at One World Community Health Centers; volunteers from the UNMC Colleges of Allied Health Professions, Pharmacy, Nursing, Dentistry, and Medicine; volunteers from Creighton University Schools of Medicine, Pharmacy, Dentistry, and College of Nursing; UNMC Global Center for Health Security program management staff; and OPS school building leadership for their efforts and dedication to the success of this project.

## FIGURE LEGENDS

**eFigure 1. Cumulative registrations and consents over the pilot program period**. Consent for testing as well as consent for treatment and ancillary clinical services was required for program participation. Student participation was limited by a high number of declinations for consent to treatment and/or ancillary clinical services.

**eFigure 2: Incremental surveillance value of asymptomatic saliva testing in the pilot program**. Number of cases identified (and so persons representing transmission risk and removed from in school activities) through the pilot in contrast to those identified by usual passive case capture.

**eFigure 3: Time interval for SARS-CoV-2 saliva PCR test conversions**.

**eFigure 4: Weekly SARS-CoV-2 case detection by saliva PCR among choir students in each pilot school**. Number of cases detected in each week is denoted in the circles.

