## Supplemental Material for "Pilot program for test-based SARS-CoV-2 screening and environmental monitoring in an urban public school district"


**eMethods**

**eTable 1.** SARS-CoV-2 RNA detection in school building air and surface samples

**eTable 2.** Demographic risk factors for SARS-CoV-2 infection in pilot participants

**eTable 3.** Acknowledgment of contributors to GISAID SARS-CoV-2 genome sequences used in this study

**eFigure 1.** Cumulative registrations and consents over the pilot program period

**eFigure 2.** Incremental surveillance value of asymptomatic saliva testing in the pilot program

**eFigure 3.** Time interval for SARS-CoV-2 saliva PCR test conversions

**eFigure 4.** Weekly SARS-CoV-2 case detection by saliva PCR among choir students in each pilot school

**eMethods**

Saliva extraction-free SARS-CoV-2 RT-PCR

The Nebraska Extraction-free Saliva (NEfS) assay used for this program was optimized for high-throughput testing with considerations for minimizing overall testing costs, overcoming potential shortages of reagents, and decreasing turnaround time. In brief, 50 µL of well-mixed saliva is added to 6.3 µL of proteinase K (New England Biolabs, P8107S), then shaken and heated at 2,200 RPM and 95°C for 5 minutes. Five microliters of the sample preparation are added to 15 µL of PCR master mix (TaqPath 1-Step RT-pPCR Master Mix, ThermoFisher Scientific A15299) containing primers and probes for detection of SARS-CoV-2 nucleoprotein RNA and human RNaseP RNA (Integrated DNA Technologies, IDT). RT-PCR is performed on QuantStudio7 Pro thermocyclers (ThermoFisher Scientific).

Waste water processing and SARS-CoV-2 RT-PCR

For RT-PCR testing, samples were processed using the Qiagen RNeasy PowerSoil Total RNA kit (Qiagen, 12866-25) following the manufacture's instructions. For each sample, 70 mL of well-mixed wastewater was divided into two 50 mL conical tubes and centrifuged at 3,500 x g for 20 minutes to concentrate solids within the specimen. The supernatant was carefully removed, and the remaining pellet was resuspended in up to 2 mL of water and used as input for the PowerSoil extraction. VetMAX™ Xeno™ Internal Positive Control RNA (ThermoFisher Scientific, A29761) was spiked into each sample to verify extraction efficiency. SARS-CoV-2 was detected using the IDT 2019-nCoV RUO kit (IDT, 10006713). A standard curve was generated using a dilution series of quantitative synthetic SARS-CoV-2 RNA (ATCC, VR-3276SD). For total and suspended solid analysis, wastewater samples were processed following ASTM D5907-18.

Air and surface sample collection and processing for SARS-CoV-2 RT-PCR

For air samples, cartridges were removed from the sampler and placed in a sealed bag for transport. Surface samples were collected using a prewetted (3 mL of sterile PBS) cotton swipe by wiping in an S-pattern, in two directions, over an approximately 100 $cm^2$ area. Following collection, surface swipes were placed into a 50 mL conical tub for transport. All air and surface samples were transported on ice to UNMC for processing. For air samples, the two metallic probes were removed from each cartridge and placed in a 15 mL conical tube with 10 mL PBS and then shaken by hand for one minute to liberate collected particles. Surface samples were recovered by adding 10 mL of PBS to the 50 mL conical tube and hand shaking for one minute. Following recovery, 400 µL of each recovered sample was extracted using the EZ1 Advanced XL Extractor (Qiagen, Hilden, Germany) with the EZ1® Virus Mini Kit v2.0

SARS-CoV-2 whole-genome sequencing

RNA was extracted from saliva samples on a KingFisher Flex (ThermoFisher Scientific) using the MagMax Viral/Pathogen II (MVP II) Nucleic Acid Isolation kit (ThermoFisher Scientific, A48383) following the manufacture's instructions for processing saliva samples. Samples were processed for sequencing by two different methods. For Oxford Nanopore Technologies (ONT) sequencing, the ARTIC multiplex PCR method V3 primer set was used following the publicly available protocol for amplification, library preparation, and analysis.[14] For Illumina-based sequencing, we used Swift Biosciences Normalase Amplicon SARS-CoV-2 V1 Panel[15] following the manufacture's instructions Libraries were sequenced on a 2 x 151 bp iSeq100 run. Genomes were assembled using the TAYLOR pipeline and consensus genomes were submitted to GISAID (EPI_ISL_1016947 – EPI_ISL_1016967).

The genomes from our study were aligned with 177 other genomes from Nebraska state, available on the GISAID repository (contributor acknowledgments provided in eTable 3) with MAFFT v1.4.0, as implemented in Geneious Prime v2021.0.3. A phylogenetic tree was inferred using RAxML v4.0 under the GTR+GAMMA model.

Statistical analyses

Data handling and analyses were performed in Microsoft Excel® and SAS®. Additional graphics were generated in GraphPad Prism 9®. In addition to descriptive epidemiology, each metadata element was assessed in univariate, and when appropriate, bivariate analyses for assessing impact on participants' first positive result. When statistically significant non-identity estimates of effect were identified or for presumed confounders that could be reasonably characterized, those elements were advanced to multivariate regression analysis. Community case rate data was not sufficiently granular to allow transforming zipcodes to levels of background COVID-19 risk. Socioeconomic status of involved zipcodes was homogenous. To limit participation effects while still exploring differential community impacts, student zip codes of residence were grouped by more to less common among students. Staff zip codes were grouped by geographic quadrant centered in downtown Omaha.

**eTable 1: SARS-CoV-2 RNA detection in school building air and surface samples**

| Pilot Week | Building Location | School A | | School B | | School C | |
|---|---|---|---|---|---|---|---|
| | | Air | Surface | Air | Surface | Air | Surface |
| **Week 1** | Choir | **P** | N | **P** | **P** | N | N |
| | Band | N | N | N | N | N | N |
| | Hallway | N | N | -- | -- | -- | -- |
| | Bathroom | -- | -- | N | N | N | N |
| | Classroom | N | N | N | N | N | N |
| | Cafeteria | N | N | N | N | N | N |
| **Week 2** | Choir | N | N | N | N | N | N |
| | Band | N | N | N | N | N | N |
| | Hallway | -- | -- | -- | -- | -- | -- |
| | Bathroom | N | N | N | N | N | N |
| | Classroom | N | N | N | N | N | N |
| | Cafeteria | N | N | N | N | N | N |
| **Week 4** | Choir | N | N | N | N | N | N |
| | Band | N | N | N | N | N | N |
| | Hallway | N | N | N | N | N | N |
| | Bathroom | -- | -- | -- | -- | -- | -- |
| | Classroom | N | N | N | N | N | N |
| | Cafeteria | N | N | N | N | N | N |
| **Week 5** | Choir | N | N | N | N | N | N |
| | Band | N | N | N | N | N | N |
| | Hallway | N | N | N | N | N | N |
| | Bathroom | -- | -- | -- | -- | -- | -- |
| | Classroom | N | N | N | N | N | N |
| | Cafeteria | N | N | N | N | N | N |

### eTable 2: Demographic risk factors for SARS-CoV-2 infection in pilot participants

| Demographics Among Students (unadjusted)[a] | | | | |
|---|---|---|---|---|
| | Overall (315) | Cases (22) | Control (293) | Association (p-value)[b] |
| Band (yes) | 62 (20) | 0 (0) | 62 (100) | 0.9503 |
| Choir (yes) | 65 (21) | 7 (32) | 58 (20) | 0.1851 |
| School | | | | |
| School A | 139 (44) | 15 (68) | 124 (42) | **0.0191** |
| School B | 71 (23) | 1 (5) | 70 (24) | |
| School C | 105 (33) | 6 (27) | 99 (34) | |
| Grade | | | | |
| 5-6 | 77 (24) | 4 (20) | 73 (25) | 0.3647 |
| 7-8 | 99 (31) | 3 (14) | 96 (33) | |
| 9-10 | 71 (23) | 7 (27) | 64 (22) | |
| 11-12 | 68 (22) | 8 (36) | 60 (20) | |

| Demographics Among Staff (unadjusted) | | | | |
|---|---|---|---|---|
| | Overall (455) | Cases (24) | Control (431) | Association (p-value) |
| School | | | | |
| School A | 178 (39) | 14 (58) | 164 (38) | 0.1528 |
| School B | 132 (29) | 5 (21) | 127 (29) | |
| School C | 145 (32) | 5 (21) | 140 (32) | |
| Position | | | | |
| Teacher | 243 (53) | 12 (50) | 231 (54) | 0.9005 |
| Administration | 58 (13) | 3 (13) | 55 (13) | |
| Cafeteria | 29 (6) | 0 (0) | 28 (7) | |
| Custodian/Security | 14 (3) | 2 (8) | 12 (3) | |
| Physical Education | 20 (4) | 1 (4) | 19 (4) | |
| Special Ed/Para | 75 (16) | 5 (21) | 70 (16) | |
| Student Support | 17 (4) | 1 (4) | 16 (4) | |

[a]Cell values reflect "N (column percent)"

[b]p-values calculated using type 3 analysis of effects

### eTable 3: Acknowledgment of contributors to GISAID SARS-CoV-2 genome sequences used in this study

We gratefully acknowledge the following Authors from the Originating laboratories responsible for obtaining the specimens, as well as the Submitting laboratories where the genome data were generated and shared via GISAID, on which this research is based.
All Submitters of data may be contacted directly via www.gisaid.org
Authors are sorted alphabetically.

| Genome entries | Originating laboratory | Submitting laboratory | Contributors |
|---|---|---|---|
| EPI_ISL_424874<br>EPI_ISL_424875 | NE Public Health Laboratory | Pathogen Discovery, Respiratory Viruses Branch, Division of Viral Diseases, Centers for Disease Control and Prevention | Yan Li, Krista Queen, Clinton R. Paden, Rachel Marine, Anna Uehara, Ying Tao, Jing Zhang, Haibin Wang, Mary S. Keckler, Alison S. Laufer Halpin, Christopher A. Elkins, Suxiang Tong |
| EPI_ISL_677521 | University of Wisconsin-Madison AIDS Vaccine Research Laboratories | University of Wisconsin-Madison AIDS Vaccine Research Laboratories | Gage Moreno, Katarina Braun, et al. AIDS Vaccine Research Laboratories |
| EPI_ISL_751563<br>EPI_ISL_751583<br>EPI_ISL_751585<br>EPI_ISL_751608<br>EPI_ISL_751638<br>EPI_ISL_751681<br>EPI_ISL_751732<br>EPI_ISL_751734 | NE Public Health Laboratory | Genomics and Discovery, Respiratory Viruses Branch, Division of Viral Diseases, Centers for Disease Control and Prevention | Krista Queen, Yan Li, Ying Tao, Jing Zhang, Anna Uehara, Anna Montmayeur, Clinton R. Paden, Peter W. Cook,Rachel Marine, Mili Sheth, Haibin Wang, Justin Lee, Suxiang Tong |
| EPI_ISL_756369<br>EPI_ISL_756370<br>EPI_ISL_756371<br>EPI_ISL_756372<br>EPI_ISL_756373<br>EPI_ISL_756374 | CUMC - CHI Bergan Mercy | Creighton University School of Medicine, Departments of Medical Microbiology and Pharmacology and Neuroscience | Michael Belshan, Morgan A. Raine, Anne V. Cheng, Christopher J. Destache, Richard V. Goering, Jacob A. Siedlik, Holly A. Stessman |
| EPI_ISL_812532<br>EPI_ISL_812550<br>EPI_ISL_812571<br>EPI_ISL_812572<br>EPI_ISL_812605<br>EPI_ISL_812627<br>EPI_ISL_812655<br>EPI_ISL_812676<br>EPI_ISL_812695<br>EPI_ISL_812705 | United States Air Force School of Aerospace Medicine | United States Air Force School of Aerospace Medicine | Anthony Fries, Jennifer Meyer, Amanda Javorina, Sarah Purves, William Gruner, Clarise Starr, Elizabeth Macias |

| | | | |
|---|---|---|---|
| EPI_ISL_831742<br>EPI_ISL_831744<br>EPI_ISL_831754<br>EPI_ISL_831819<br>EPI_ISL_831825<br>EPI_ISL_831834<br>EPI_ISL_831841<br>EPI_ISL_831857<br>EPI_ISL_831866<br>EPI_ISL_831887 | United States Air Force School of Aerospace Medicine | United States Air Force School of Aerospace Medicine | Anthony Fries, Jennifer Meyer, William Gruner, Amanda Javorina, Sarah Purves, Clarise Starr, Elizabeth Macias |
| EPI_ISL_850933 | Helix / Illumina | Genomics and Discovery, Respiratory Viruses Branch, Division of Viral Diseases, Centers for Disease Control and Prevention | Peter W. Cook, Dhwani Batra, Ben L. Rambo-Martin Eileen de Feo, Jan Antico, Christine Tran, Matthew Tolentino, Shannon Wickline, Kim Gietzen, Brad Sickler, Jingtao Liu, Eric Allen, Phil Febbo, Summer Galloway, Nicole L. Washington, Simon White, Geraint Levan, Kelly Schiabor Barrett, Elizabeth Cirulli, Alexandre Bolze, Ary Ascencio, Charlotte Rivera-Garcia, Ryan Cho, Jason Nguyen, Sherry Wang, Jimmy Ramirez, Tyler Cassens, Efren Sandoval, Magnus Isaksson, William Lee, David Becker, Marc Laurent, James Lu, Clinton R. Paden, Suxiang Tong, Duncan MacCannell |
| EPI_ISL_876873 | Quest Diagnostics | Quest Diagnostics | Rosenthal,S.H., Gerasimova,A., Kagan,R.M., Anderson, B., Hua, M., Liu Y., Bernstein, L.E., Livingston, K.E., Perez, A., Shalhout, D.F., Shlyakhter, I.A., Owen, R., Tanpaiboon, P., Lacbawan, F. |
| EPI_ISL_884375 | Infectious Diseases, Quest Diagnostics | Infectious Diseases, Quest Diagnostics | Rosenthal,S.H., Gerasimova,A., Kagan,R.M., Anderson,B., Bernstein,L.E., Livingston,K.E., Hua,M., Liu,Y., Shalhout,D.F., Owen,R., Lacbawan,F. |
| EPI_ISL_886189<br>EPI_ISL_886258<br>EPI_ISL_886278<br>EPI_ISL_886334<br>EPI_ISL_886376<br>EPI_ISL_886377<br>EPI_ISL_886430<br>EPI_ISL_886530<br>EPI_ISL_886544<br>EPI_ISL_886596<br>EPI_ISL_886620<br>EPI_ISL_886696<br>EPI_ISL_886713<br>EPI_ISL_886766<br>EPI_ISL_886786<br>EPI_ISL_886787<br>EPI_ISL_886826<br>EPI_ISL_886908<br>EPI_ISL_886958<br>EPI_ISL_887096<br>EPI_ISL_887699<br>EPI_ISL_888372<br>EPI_ISL_888468 | Labcorp | Genomics and Discovery, Respiratory Viruses Branch, Division of Viral Diseases, Centers for Disease Control and Prevention | Peter W. Cook,Dhwani Batra,Ben L. Rambo-Martin,Summer Galloway,Brian Krueger,Minoo Agarwal,Eyad Almasri,Debbie Boles,Ayla Burns,Nuthawin Charoensri,Oren Cohen,Susan Countryman,Mary Ann Cristobal,Bobbi Croy,Suzanne Dale,Hrushikesh Deshmukh,Amanda Douglas,Vincent Drouillon,Marcia Eisenberg,Howard Engler,Rama Ghatti,Prashant Gupta,Susan Hicks,Jake Humphrey,Lax Iyer,Manoj Jain,Mohan Kolli,Tim Kuphal,Stanley Letovsky,Michael Levandoski,Craig Lukasik,Jonathan Meltzer,Brian Norvell,Mindy Nye,Scott Parker,Christos Petropoulos,John Pruitt,Steven Ragan,Scott Ryan,Mike Sapeta,Jana Schroth,Suresh Babu Selvaraju,Goran Stevovic,Amanda Suchanek,Andrea Throop,Lyndon Tilson,Thomas Urban,Joe Voshell,Kimberly Wagner,Jonathan Williams,Mary Williamson,Qian Zeng,Tricia Zwiefelhofer,Clinton R. Paden,Suxiang Tong,Duncan MacCannell, |
| EPI_ISL_903747<br>EPI_ISL_903774<br>EPI_ISL_903791<br>EPI_ISL_903817<br>EPI_ISL_903845<br>EPI_ISL_903885<br>EPI_ISL_903901 | NE Public Health Laboratory | Genomics and Discovery, Respiratory Viruses Branch, Division of Viral Diseases, Centers for Disease Control and Prevention | Krista Queen, Yan Li, Ying Tao, Jing Zhang, Anna Uehara, Anna Montmayeur, Clinton R. Paden, Peter W. Cook, Rachel Marine, Mili Sheth, Jasmine Padilla, Sarah Nobles, Mark Burroughs, Lori Rowe, Haibin Wang, Ben L. Rambo-Martin, Dhwani Batra, Justin Lee, Suxiang Tong |

| | | | |
|---|---|---|---|
| EPI_ISL_428202 | Nebraska Public Health Laboratory | UNMC COVID-19 Response Team | UNMC COVID-19 Response Team |
| EPI_ISL_428203 | | | |
| EPI_ISL_428204 | | | |
| EPI_ISL_428205 | | | |
| EPI_ISL_428206 | | | |
| EPI_ISL_466652 | | | |
| EPI_ISL_466653 | | | |
| EPI_ISL_466654 | | | |
| EPI_ISL_466655 | | | |
| EPI_ISL_466656 | | | |
| EPI_ISL_466657 | | | |
| EPI_ISL_466658 | | | |
| EPI_ISL_466659 | | | |
| EPI_ISL_466660 | | | |
| EPI_ISL_466661 | | | |
| EPI_ISL_466662 | | | |
| EPI_ISL_466663 | | | |
| EPI_ISL_466664 | | | |
| EPI_ISL_466665 | | | |
| EPI_ISL_466666 | | | |
| EPI_ISL_466667 | | | |
| EPI_ISL_466668 | | | |
| EPI_ISL_466669 | | | |
| EPI_ISL_466670 | | | |
| EPI_ISL_466671 | | | |
| EPI_ISL_466672 | | | |
| EPI_ISL_466673 | | | |
| EPI_ISL_466674 | | | |
| EPI_ISL_466675 | | | |
| EPI_ISL_466676 | | | |
| EPI_ISL_466677 | | | |
| EPI_ISL_466678 | | | |
| EPI_ISL_466679 | | | |
| EPI_ISL_466680 | | | |
| EPI_ISL_466681 | | | |
| EPI_ISL_466682 | | | |
| EPI_ISL_466683 | | | |
| EPI_ISL_467298 | | | |
| EPI_ISL_475174 | | | |
| EPI_ISL_475175 | | | |
| EPI_ISL_475176 | | | |
| EPI_ISL_475177 | | | |
| EPI_ISL_475178 | | | |
| EPI_ISL_475179 | | | |
| EPI_ISL_475180 | | | |
| EPI_ISL_475181 | | | |
| EPI_ISL_475182 | | | |
| EPI_ISL_475183 | | | |
| EPI_ISL_475184 | | | |
| EPI_ISL_475185 | | | |
| EPI_ISL_475186 | | | |
| EPI_ISL_475187 | | | |
| EPI_ISL_475188 | | | |
| EPI_ISL_475189 | | | |
| EPI_ISL_475191 | | | |
| EPI_ISL_475192 | | | |
| EPI_ISL_475193 | | | |
| EPI_ISL_475194 | | | |
| EPI_ISL_475195 | | | |
| EPI_ISL_475196 | | | |
| EPI_ISL_475197 | | | |
| EPI_ISL_475198 | | | |
| EPI_ISL_475199 | | | |
| EPI_ISL_475200 | | | |
| EPI_ISL_475201 | | | |
| EPI_ISL_475202 | | | |
| EPI_ISL_475203 | | | |
| EPI_ISL_475204 | | | |
| EPI_ISL_475205 | | | |
| EPI_ISL_475206 | | | |

| |
|---|
| EPI_ISL_475207 |
| EPI_ISL_475208 |
| EPI_ISL_475209 |
| EPI_ISL_475210 |
| EPI_ISL_475211 |
| EPI_ISL_475212 |
| EPI_ISL_475213 |
| EPI_ISL_475214 |
| EPI_ISL_475215 |
| EPI_ISL_475216 |
| EPI_ISL_475217 |
| EPI_ISL_475218 |
| EPI_ISL_475219 |
| EPI_ISL_475220 |
| EPI_ISL_475221 |
| EPI_ISL_475222 |
| EPI_ISL_475223 |
| EPI_ISL_475224 |
| EPI_ISL_475225 |
| EPI_ISL_475226 |
| EPI_ISL_475227 |
| EPI_ISL_475228 |
| EPI_ISL_475229 |
| EPI_ISL_475230 |
| EPI_ISL_475231 |
| EPI_ISL_475233 |
| EPI_ISL_475234 |
| EPI_ISL_475235 |
| EPI_ISL_475236 |
| EPI_ISL_475237 |
| EPI_ISL_732820 |
| EPI_ISL_732821 |
| EPI_ISL_732822 |
| EPI_ISL_732823 |
| EPI_ISL_732824 |
| EPI_ISL_732825 |
| EPI_ISL_732826 |

**eFigure 1: Cumulative registrations and consents over the pilot program period.** Consent for testing as well as consent for treatment and ancillary clinical services was required for program participation. Student participation was limited by a high number of declinations for consent to treatment and/or ancillary clinical services.

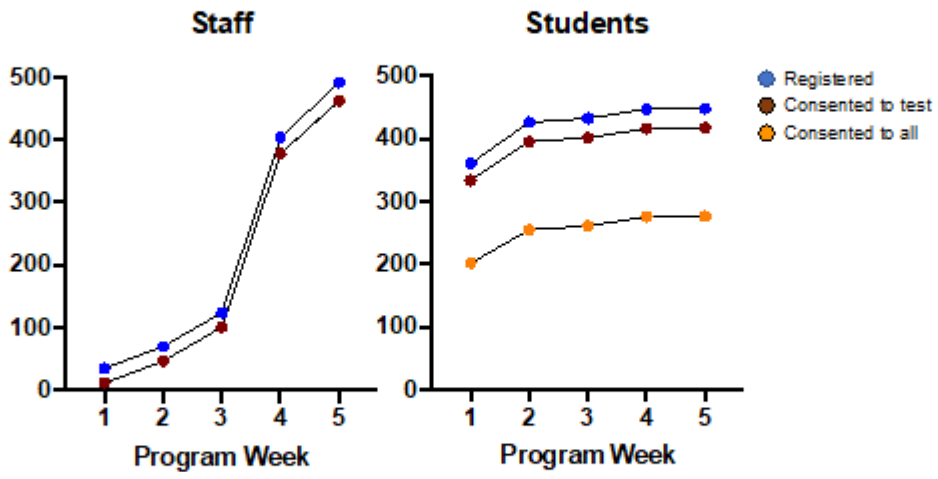

**eFigure 2: Incremental surveillance value of asymptomatic saliva testing in the pilot program.**
Number of cases identified (and so persons representing transmission risk and removed from in school activities) through the pilot in contrast to those identified by usual passive case capture.

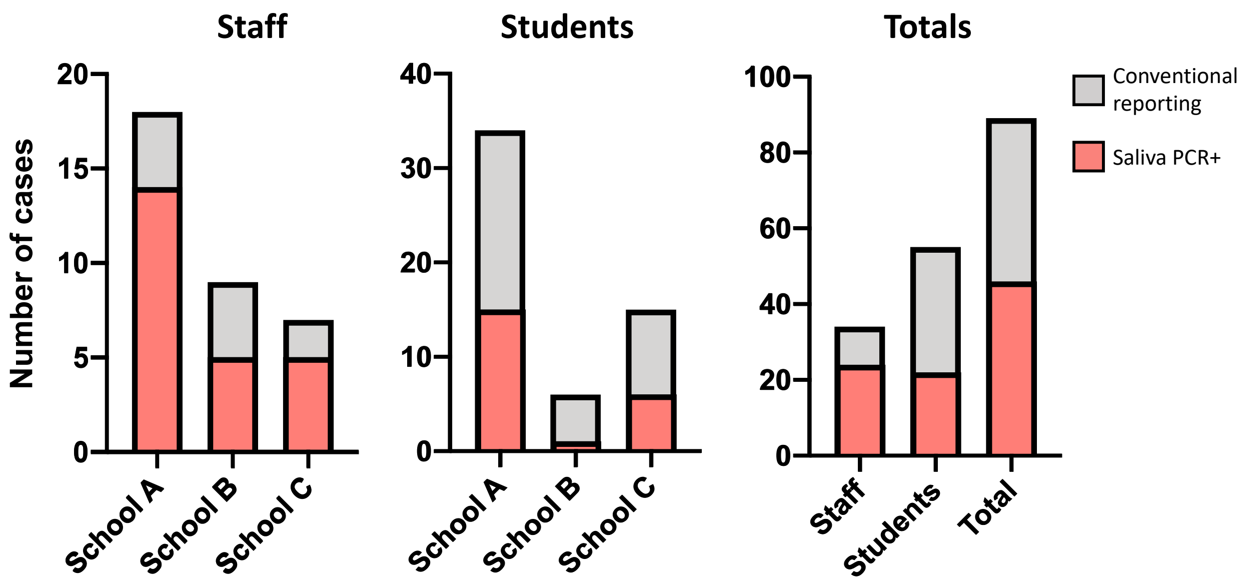

**eFigure 3: Time interval for SARS-CoV-2 saliva PCR test conversions.**

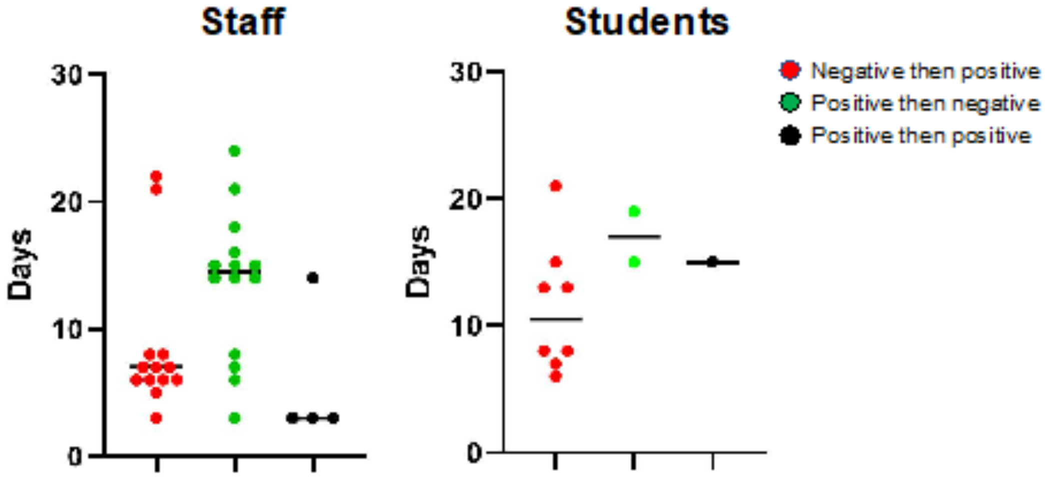

**eFigure 4: Weekly SARS-CoV-2 case detection by saliva PCR among choir students in each pilot school.**
Number of cases detected in each week is denoted in the circles.

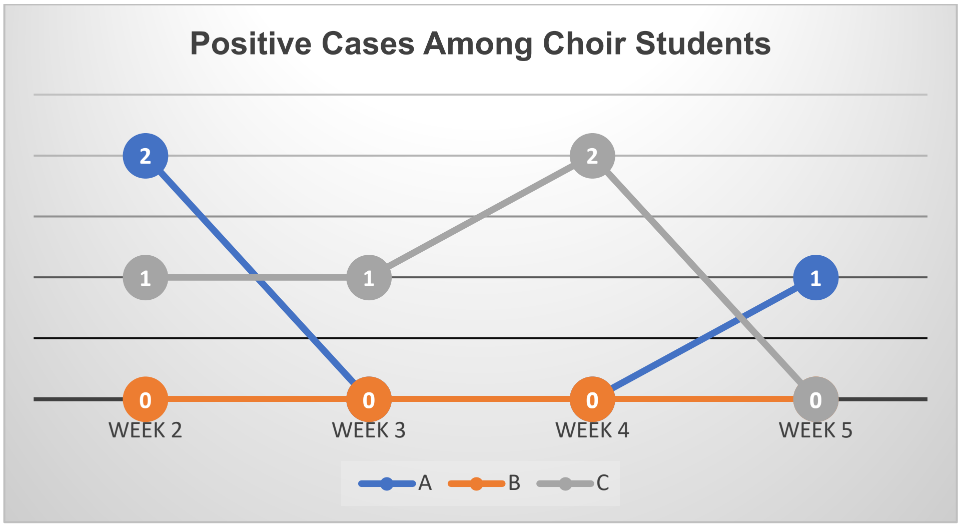
